# An unsupervised deep clustering for Bone x-ray classification and anomaly detection

**DOI:** 10.1101/2023.04.16.23288653

**Authors:** Guola Zhou, Caiping Hu, Yulin Zhang, Jieru Jiang

## Abstract

In the medical field, bone abnormality detection is a very important issue. Bone abnormalities include various diseases such as fractures, osteoporosis, bone tumors, and joint diseases. If these diseases are not diagnosed and treated in a timely manner, they can seriously affect the health and quality of life of patients. Artificial intelligence has made remarkable advances in Cluster analysis of medical big data, effectively mining its hidden associations to provide effective information for clinical diagnosis and medical research. However, the effectiveness of deep learning in domains with limited or no labeled data is often limited. To address this issue, we propose a novel and reliable two-stage unsupervised deep clustering framework for skeletal anomaly detection. This framework combines neural network parameters with feature clustering for collaborative learning to detect anomalies. We trained eight separate models, one for classification and seven for anomaly detection, using the MURA dataset, the largest publicly available skeletal imaging dataset. In the first stage, our approach achieved an average sensitivity and specificity of 99.76% and 99.53%, respectively. The second stage performed optimally with an average sensitivity and specificity of 83.28% and 97.56%, respectively. Our method can be easily implemented as software modules and used as a visualization tool for skeletal physicians, making it a promising approach for future development.

## 1. Introduction

Bone classification and abnormality detection are of great significance in the medical field. Bone classification refers to the classification of bone structure and morphology to better understand and study the physiological and pathological state of bones[1]. At the same time, bone abnormality detection is aimed at the diagnosis and monitoring of bone diseases, in order to better improve the treatment effect and quality of life of patients [2]. According to a study, the accuracy of bone abnormality detection has an important impact on the treatment effectiveness and prognosis of patients. This study indicates that early diagnosis and treatment of skeletal abnormalities can greatly improve the prognosis of patients. Accurate detection of bone abnormalities can enable doctors to identify conditions faster and provide better treatment options [3]. In addition, bone abnormality detection can also help doctors better understand the development and progress of bone diseases [4]. A study from the National Institutes of Health in the United States indicates that through in-depth understanding of the classification and abnormality detection of skeletal diseases, it is possible to better understand the progress of the disease and adjust and optimize treatment plans.

In recent years, the application of machine learning has become increasingly widespread, with traditional clustering being the most typical, which can automatically complete classification and detection tasks without human intervention. Clustering is an unsupervised learning algorithm whose main purpose is to divide data points into different groups or clusters based on their similarity [5]. Clustering algorithms have significant applications in many fields, such as: in data mining: clustering can help us discover important patterns and trends in big data sets, thereby making data mining more efficient and accurate [6]; In biology: clustering can be used to classify and analyze gene sequences, thereby helping scientists better understand the complexity of genetics and biology [7]; Computer Vision: Clustering can be used to classify and analyze images [8]. For example, we can use clustering algorithms to automatically identify different objects in an image [9]. In addition, clustering can be used for text classification tasks [10], such as dividing a group of text data into multiple categories, so that texts within the same category have similar thematic, emotional, and other characteristics [11]. In the medical field, traditional clustering can classify and predict diseases, but clustering algorithms have high requirements for the quality of data. If there are problems such as noise or missing values in the data, it may lead to inaccurate clustering results, which affects the reliability of medical decision-making. In addition, deep learning can be applied to digital pathology to complete automatic diagnosis. For example, techniques such as clinically assisted diagnosis [12], tumor segmentation [13], and protein structure prediction [14]. However, the effectiveness of a deep learning framework largely depends on the training data and the number of valid tags, because: whether to choose an appropriate loss function, small batch size, learning rate, and so on. For fields with few or no labels, there are some existing methods: semi supervised learning [15], weak supervision [16], and domain adaptation [17]. Although some existing studies have been applied to the diagnosis of supervised bone diseases, it is still challenging to effectively apply unsupervised frameworks to the rapid clinical diagnosis of bone diseases. Although good progress has not been achieved in traditional clustering, in this study, we have developed a deep clustering framework, which is different from the previous fully supervised learning framework. Its effectiveness depends on a large amount of training data and tags. By combining deep learning, the clustering of neural network parameters and related features is allocated together for joint learning. The k-means clustering algorithm is used to iteratively group features, and then the resulting groups are used as supervised pseudo tags to update the weight of the network. The framework method is used to iteratively learn features, group them, and update network parameters. In this article, we apply the MURA (Musculoskeletal Radiology) dataset, which is a large dataset of bone X-rays and has seven main categories, including ELBOW, FINGER, FOREARM, HAND, HUMERUS, SHOULDER, and WRIST. A total of 36487 images are included. The experimental results show that the optimal average sensitivity and specificity for the first stage are 99.76% and 99.53%, respectively. The optimal average sensitivity and specificity for the second stage were 83.28% and 97.56%, respectively. Finally, this framework greatly affects the future assistance of doctors, and has a good application prospect.

Our contributions can be summarized as follows:

1. Deep clustering is an unsupervised end-to-end learning method clustering algorithm applicable to any standard convective network. The framework is to jointly learn the parameters of the neural network and the clustering assignment of the result characteristics. Feature extraction is performed using convnet, features are learned iteratively and grouped using Deep Clustering method and subsequent results are used as supervised pseudo-labels to update the network weights.
2. In this paper, we applied the MURA dataset, a large dataset of skeletal X-rays, with seven major categories: ELBOW, FINGER, FOREARM, HAND, HUMERUS, SHOULDER, WRIST. a total of 36487 images are included.
3. The experimental findings showed that the first stage’s optimal average sensitivity and specificity were 99.76% and 99.53%, respectively. The optimal average sensitivity and specificity were 83.28% and 97.56%, respectively, during the second phase. It demonstrates that the framework has good and significant results and has good prospects for the future development.

The remainder of this essay is structured as follows: Basic ideas are introduced in Section 2, the suggested clustering method is described in Section 3, and the experimental results of the proposed algorithms on diverse data sets are shown in Section 4. The conclusion and our next efforts are provided in Section 5.

## 2. Related Work

### 2.1 Deep clustering

According to traditional clustering, we are familiar with algorithms such as DBSCAN, K-means, etc. The DBSCAN algorithm requires two parameters: the scan radius ε, and the MinPts, which is used to determine the density threshold of points being core points. The core objective of the K-means algorithm is to divide the given data set into K clusters (K is the hyperparameter) and give each sample data corresponding centroid. As mentioned above, therefore, similar to clustering, these methods are very complex and time-consuming. On this basis, Li et al. [18] proposed an improved DBSCAN based on neighbor similarity, which significantly and effectively improved the efficiency of the DBSCAN algorithm; Chen et al. [19] proposed the BLOCK-DBSCAN targeted to deal with high-dimensional datasets and made a breakthrough in effect; Lika et al. [20] proposed the global k-means algorithm and a modification of this method was tested on numerous datasets and obtained better results. Lai et al. [21] proposed a fast mean clustering algorithm (FKMCUCD) that can effectively reduce the computation time. Although all such algorithms can solve the corresponding problems at some level and improve the efficiency of the algorithm, this class of algorithms cannot be applied to other technical levels, especially at the image level.

The advancement of deep learning has resulted in incredible advancements in numerous industries, especially in images, where the accuracy has reached a level that exceeds that of humans. Based on different distance metrics, Peng et al. [22] will lead to similar soft clustering assignments on stream shapes. A new clustering method is proposed: one of the first end-to-end approaches to jointly learn clustering assignments for clustering methods solving image-level problems. In order to implicitly anticipate segmentation labels of target spectral maps from the input mixture, Hershey et al. [23] develop discriminative embedding of segmentation and separation in a deep learning framework termed deep clustering. A deep network-based simulation of spectral clustering results from this. AGC IMC, or Adaptive Graph Completion-based Incomplete Multiview Clustering, was proposed by Wen et al. [24]. AGC IMC greatly outperforms other cutting-edge techniques for graph completion and consensus representation learning, which was developed together. Zhang et al. [25] proposed a fast and efficient CNN (convolutional neural network) denoiser that performs well for various low-level vision applications. In order to advance the area of image segmentation, Ghosh et al. [26] discussed the significance of deep learning for image segmentation and how it has affected the field. To address the issue of picture clustering, Ren et al. [27] suggested a two-stage depth density based image clustering (DDC) framework. In comparison to cutting-edge deep clustering techniques, the suggested DDC delivers similar or superior clustering performance. All of the above methods can solve specific problems in specific domains and all have particularly good results. However, no previous work has been done for the time being to explore their application in solving skeletal x-ray image recognition, so it is still a challenge here.

### 2.2 Intelligent recognition of skeletal data

Image classification is one of the most in-demand technologies of our time and is used in various fields such as healthcare, business, etc. Giving an image a label from a list of classifications is the essence of image categorization. In actuality, this implies that our job is to examine an input image and provide a label that will assign the image to one of a predetermined number of possible categories. In traditional methods, we usually solve the image classification problem using two methods, k-NN (k-Nearest Neighbor) and Support Vector Machine (SVM). For example, the complete dataset was divided into parts by Zhen et al. [28] using k-means clustering, and each component was subsequently subjected to k-NN classification. A series of experimental imaging data was performed in big data and medicine. One of the simplest image categorization techniques available is k-NN. It can be used in regression analysis as well. Since k-NN is a parameter-free learning algorithm, no assumptions are made on the distribution of the underlying data. It is instance-based, i.e. the algorithm does not consciously learn the model. Instead, it chooses memory training examples and applies them in a controlled learning setting. The three essential components of the k-NN algorithm are the decision rule for classification, the k-value selection, and the choice of distance computation. The most frequent class among the k nearest examples is determined using the k-NN algorithm, which categorizes unknown data points. We must establish a distance metric or similarity function before we can use k-nearest neighbor classification. The Manhattan distance and Euclidean distance are popular options. Li et al. [29] proposed a multi-labeled SVM active learning method. Two selection strategies are proposed: the maximum loss strategy and the average maximum loss strategy. A linear classifier constructed by interval maximization on the feature space is the basis of the SVM, a binary classification model. It differs from a perceptron thanks to interval maximization, and SVM also incorporates the kernel method, effectively turning it into a nonlinear classifier. Interval maximization, which is formalized as problem solving, is the SVM’s learning technique. Convex quadratic programming is solved using the optimization procedure of the SVM. Finally, this traditional method has shortcomings: it is only suitable for small datasets, and the results are slightly worse for large datasets.

### 2.3 Deep Learning Methods

Convolutional neural network is a deep learning model widely used in the fields of image processing and computer vision. The principle is to extract features from input images through convolution and pooling operations, and then perform tasks such as classification or regression through the full connectivity layer. Additionally, it is a feedforward neural network with artificial neurons that can analyze enormous amounts of data and react to nearby units. To effectively complete image classification tasks, Sun [30] suggested an automated CNN architecture design technique using evolutionary algorithms. The algorithm uses fewer computational resources and is effective. Finally, classical algorithms like SVM are insufficient for picture categorization when compared to KNN. Even with overfitting in CNN, the experimental results are superior to those of conventional classification methods. When it comes to image categorization issues, transfer learning works incredibly well. It effectively addresses the issues of overfitting and tiny datasets thanks to its quick running time and precise findings. In the previous work are exploring the fully supervised framework, it is still a challenge to apply the unsupervised framework to skeletal x-ray image recognition effectively.

## 3. Methodology

This paper proposes a brand-new, two-stage hybrid approach based on deep clustering for the classification and anomaly detection of skeletal x-rays. As shown in Fig 1, the original images are first trained by a neural network to obtain a training dataset, which is preprocessed, and important features are extracted. The retrieved features are then used in a two-stage classification process. First, the seven skeleton types—ELBOW, FINGER, FOREARM, HAND, HUMERUS, SHOULDER, and WRIST—are classified. The second stage of the unsupervised deep clustering framework detection technique is to determine if the classified skeletons are normal or abnormal, as shown in Fig 2. All the necessary information is provided in the following subsections:

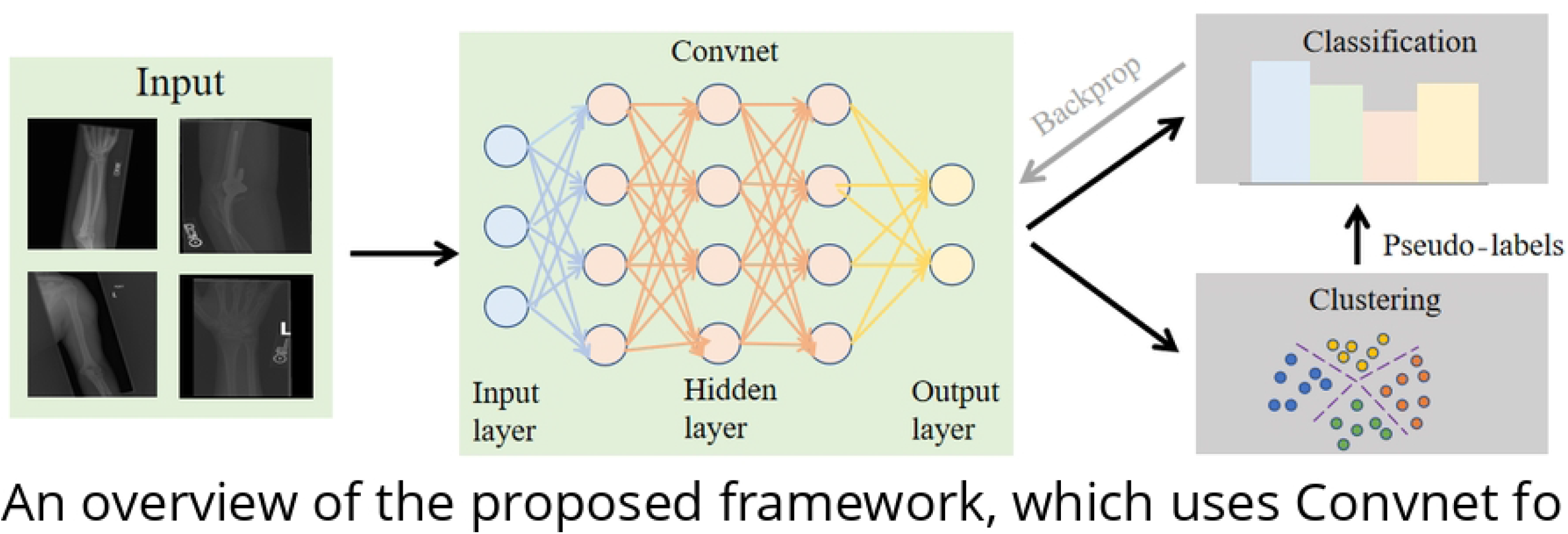

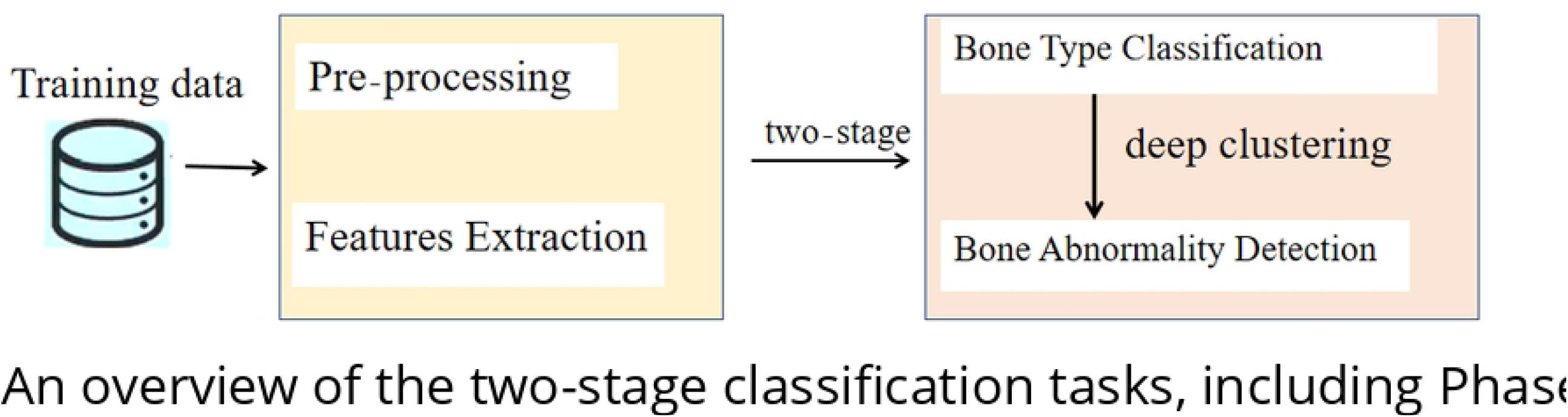

### 3.1. Preprocessing and feature extraction

The current computer vision outlook should have good image extraction characterization and convolutional neural network architecture (Convnet) has significantly impacted computer vision research. A convolutional network basically learns a large number of mapping relationships between input and output for mapping the original image to a vector space of fixed dimensions. As long as the convolutional network is trained with recognized patterns, it may map between input and output pairs without needing an exact mathematical expression between input and output. An input image is converted into a matrix, which contains the appropriate pixel values, for that image. A feature map is obtained by convolution using a convolution kernel.

#### 3.1.1 Local perception and Parameter sharing

Just the local information is actually required for each neuron to perceive, and the global information is then gained by merging the local information at a higher level. In local connectivity, as a result of each neuron in the hidden layer being connected to a 10 by 10 local image, there are 10 by 10 weight parameters. The remaining neurons share these 10 by 10 weight parameters, making all of the neurons in the hidden layer have the same weight parameters. Then, regardless of the quantity of neurons in the hidden layer, the parameters that must be learnt are these 10 by 10 weight parameters. This is the convolution kernel’s size. therefore, the statistical characteristics of one component of the image are the same as those of the other components.

#### 3.1.2 Convolution operations

We initially number each pixel in the image by designating the i-th row and j-th column parts of the image in order to clearly understand the convolution calculation method; By indicating the m-th row and n-th column weights for each filter weight and the bias term for the filter, the weights are numbered; The activation function is represented by the f element of the Feature Map. The activation function is represented by the i-th row and j-th column elements of the Feature Map (the relu function is chosen as the activation function for this example). The following equation is then used to calculate the convolution:

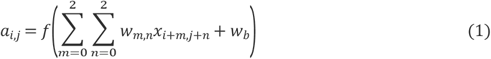

Additionally, its picture size, step size, and convolved Feature Map size are connected. They in fact fulfill the following relationship:

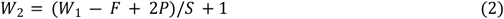

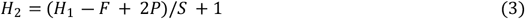

*W*_1_is the width of the picture before convolution, *W*_2_ is the width of the Feature Map after convolution, and F is the width of the filter in Equations 2 and 3. P is the number of Zero Padding, or the number of circles surrounding the original image that are zero; if P is 1, then there will be one circle that is zero; S is the step size; *H*_1_ is the width of the picture before to convolution, and *H*_2_ is the height of the Feature Map.

In fact, it is similar. The depth of the corresponding filter must likewise be D if the depth of the picture before convolution is D. We extend Equations 1 to obtain the formula for convolution with a depth greater than 1.

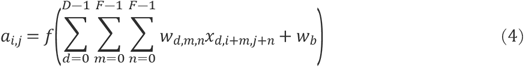

In Equation 4, D is the depth; F is the size of the filter; denotes the m-th row and nth column weight of the d-th layer of the filter; denotes the i-th row and j-th column pixel of the d-th layer of the image; the other symbols have the same meaning as Equation 1. There may be several filters and a for each convolution layer. The depth (number) of the convolved Feature Map and the number of filters in the convolution layer are equal because a Feature Map can be created by convolving each filter with the original picture.

#### 3.1.3 Pooling and BP

We know how CNN uses convolutional and pooling layers to extract features from images, where the key is the convolutional kernel to represent the local features in the image. This section introduces backpropagation, which is the process of contrasting the anticipated value with the actual value before returning to adjust the network parameters. Also the operation of the backpropagation algorithm is described here: by first calculating the state and activation values for each layer up to the last layer, then calculating the error for each layer, the error is calculated by moving forward from the last layer and finally updating the parameters, i.e. the process of comparing the expected value with the actual value before returning to adjust the network parameters.

This section mainly explains the principle process of basic CNN. The general CNN technique for extracting features uses the convolutional layer, the pooling layer, and back propagation to calculate the convolutional kernel parameters in order to obtain the final features.

### 3.2 Unsupervised Two-Stage classification and detection

These parameters are typically acquired through supervision, meaning that each image, *x*_*n*_, has a corresponding label, *y*_*n*_, in the range of 0 to 1k. This label indicates how the image fits into one of the k predefined classes that are possible. Over the feature f, the parameterized classifier *g*_*w*_ forecasts the correct label (*x*_*n*_). The loss function is as a result.

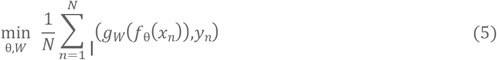

where the gradient is calculated using small-batch stochastic gradient descent and backpropagation to minimize this loss function, and *ℓ* is the multinomial logarithmic loss. In order to improve the previously given algorithm, we cluster the output of convnet and use the outcomes of the subsequent clustering as “pseudo-labels.” This deep clustering method iteratively learns features and groups them. Where clustering is done using the standard clustering algorithm k-means.

K-means clustering accepts a set of input vectors and clusters the features that are displayed. from the convolutional neural network (n features representing n images) into k classes. Then it assigns a pseudo-label y to each sample x according to the above loss function, which is actually k-means clustering.

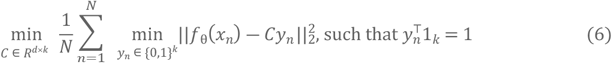

Using this formula to jointly learn the d × k prime matrix C and the clustering assignment *y*_*n*_ for each image n, overall, deep cluster alternatively clusters the features using Equations (6) to generate pseudo labels or updates the parameters of the network by predicting these pseudo labels using Equations (5). The deep clustering method introduced in Equation (6) has been shown to improve the performance of traditional supervised learning methods by leveraging the unsupervised nature of clustering algorithms to learn more informative feature representations. This approach is particularly effective when labeled data is scarce or expensive to obtain, as it allows the model to learn from the vast amounts of unlabeled data that are available.

One advantage of deep clustering is that it can be used to learn representations that are more robust to variations in the input data, such as changes in lighting or perspective. By clustering the features of multiple images, the model can identify patterns that are consistent across different instances of the same object or scene, and use this information to improve its ability to classify new images.

Another benefit of deep clustering is that it can be used to perform unsupervised feature learning, which can be used to pretrain the parameters of a neural network before fine-tuning it on a smaller labeled dataset. This can help to mitigate the problem of overfitting, which can occur when the model is trained on a limited amount of labeled data. By pretraining the model on a larger amount of unlabeled data, the model can learn more generalizable feature representations that can be fine-tuned to specific tasks with smaller amounts of labeled data.

Overall, deep clustering is a promising approach to improve the performance of supervised learning methods, and has been shown to be effective in a wide range of computer vision tasks, including object recognition, image retrieval, and semantic segmentation. By leveraging the complementary strengths of supervised and unsupervised learning, deep clustering can help to overcome the limitations of traditional supervised learning methods and unlock new possibilities in computer vision research.

### 3.3 Avoiding tractable solutions

When using clustering, one encounters the case of a nontrivial solution. A nontrivial solution is a network that, if not qualified during the training process, may take shortcuts to fit the loss function and learn nothing instead. Two options to avoid this are mentioned in this paper.

#### 3.3.1 Empty clusters

Using the model to predict pseudo-labels may cause the features generated by the network to be clustered around a certain cluster center, leaving other cluster centers without samples, this is because there is no restriction that a certain cluster center cannot have no samples. When a cluster center is empty, one option is to randomly choose a non-empty cluster center and add some small perturbations to it as a new cluster center while letting the samples belonging to the non-empty cluster center also belong to the new cluster center. However, this requires computing the entire dataset and is too expensive. Another option is to limit the minimum number of samples per cluster center.

#### 3.3.2 Trivial parametrization

The fact that a lot of data is grouped into a limited number of classes is another issue. To be grouped into a single class represents an extreme case in which the network might result in the same output for any given set of inputs. The answer to this issue is to uniformly sample the samples depending on categories (or pseudo-labels). This approach is also equivalent to weighting the loss function according to the number of Clusters in each.

In this paper, we use a two-stage unsupervised clustering approach where the deep clustering framework utilizes convnet for preprocessing and feature extraction. The output is clustered and the results of the subsequent clustering are used as “pseudo-labels”, which are iteratively learned and grouped using deep clustering methods. Finally, anomaly detection is performed by clustering and classification.

## 4. Experimental Results

The previous section was used to input the unsupervised neural network in order to extract the most important features. After then, the features were divided into two phases. ELBOW, FINGER, FOREARM, HAND, HUMERUS, SHOULDER, and WRIST were the first seven bone types assigned to the input bone X-rays. Then, the second stage detects abnormalities in that bone type by training the classifier with normal or abnormal skeletal measurements. The next subsection provides information about: 1. the utilized dataset; 2. the experimental details; and 3. the experimental results.

### 4.1 dataset

This study used the MURA dataset, one of the largest publicly available skeletal imaging databases, for training and testing. There are seven main categories, totaling 9067 normal and 5915 pathological musculoskeletal rays in the upper extremity: the ELBOW, FINGER, FOREARM, HAND, HUMERUS, SHOULDER, WRIST. Each research includes several skeleton perspectives. Consequently, there are 40561 multi-view X-ray images in the MURA database. There are three main groups are present:

1. training (11,255 patients, 13,565 studies, 37,111 images)
2. Reliable (788 patients, 1208 studies, 3225 images)
3. Examining (208 patients, 209 studies, 559 images)

MURA is a musculoskeletal radiograph dataset containing 40561 multi-view radiographic images from a total of 14863 studies in 12173 patients. Each one falls into one of seven standard upper extremity radiology study types. Each one fits into one of these categories. From 2001 through 2012, a board-certified radiologist from the Stanford School of Medicine manually classified each study as normal or abnormal depending on how diagnostic radiology interpreted clinical medical imaging.

To evaluate the model and obtain radiologist-level robustness estimates, the investigators obtained additional labeling from six Stanford radiologists with board certification, including 207 musculoskeletal studies. Radiologists had an average of 8.32 years of practice experience, with years of practice ranging from 2 to 25. The investigators randomly selected 3 radiologists to construct the golden rule, which was defined as the label voted on by the majority of radiologists.

In this paper, the experimental set has been performed in terms of training and validation. The amount of samples per bone utilized for testing and training is displayed in Tab 1.

### 4.2 Implementation details

For the classification and anomaly detection of skeletal x-rays, an unique two-stage hybrid technique based on deep clustering is put forth in this study. The network framework used is ResNet50, which uses a residual structure to solve the problem of deep network degradation and difficulty in training for a variety of computer vision tasks by enabling the network to be deeper, converge faster, and optimize more easily while having fewer parameters and lower complexity compared to previous models.

All experiments were performed on an Intel Core(TM) i5-12600K CPU with 6 cores, 16 GBRAM and RTX 3070. PyTorch version 1.4.0, pickleshare package 0.7.5, pillow 6.2.1, scikit -image is 0.15.0 and scikit-learn is 1.0.2.

### 4.3 Experimental results

Sensitivity, specificity, are the two-stage methodologies used to gauge the effectiveness of the suggested procedure. The formulae underlying these measurements, which are based on using the actual normalized moments of confusion, are displayed below.

Authors should discuss the results and how they can be interpreted from the perspective of previous studies and of the working hypotheses. The findings and their implications should be discussed in the broadest context possible. Future research directions may also be highlighted.

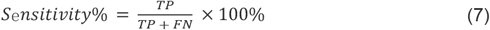

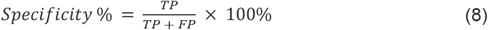

where TP is the number of cases from the specified class that are correctly classified and FN is the number of examples from the given class that are wrongly classified as belonging to another class; The percentage of correctly identified instances of each skeleton (class) indicates how accurate it is; where FP is the number of instances from other classes that are incorrectly categorized as belonging to the given class (seven skeleton types) which passes through a series of convolution operations. Subsequently, the pre-processed images were sent to different pre-trained CNN model layers, and these were examined separately for the two stages. For stage 1, as shown in Table 2, the results show an optimal average sensitivity and specificity of 99.76% and 99.53%, respectively.

**Tab 1.** The training and testing number of samples used in the experiments

| Bone Type | ELBOW | FINGER | FOREARM | HAND | HUMERUS | SHOULDER | WRIST |
| --- | --- | --- | --- | --- | --- | --- | --- |
| Testing | 4931 | 5106 | 1825 | 5543 | 1272 | 8379 | 9752 |
| Training | 4931 | 5106 | 1825 | 5543 | 1272 | 8379 | 9752 |

**Tab. 2.** Sensitivity and Specificity of deep clustering model for Stage 1

| Bone | ELBOW | FINGER | FOREARM | HAND | HUMERUS | SHOULDER | WRIST | AVERAGE |
| --- | --- | --- | --- | --- | --- | --- | --- | --- |
| Sensitivity | 99.56% | 99.97% | 99.54% | 99.58% | 99.92% | 99.86% | 99.89% | 99.76% |
| Specificity | 99.56% | 99.25% | 99.67% | 99.78% | 99.79% | 99.49% | 99.17% | 99.53% |

**Tab 3.** Sensitivity and Specificity results of deep clustering models in Stage 2

| Bone | ELBOW | FINGER | FOREARM | HAND | HUMERUS | SHOULDER | WRIST | AVERAGE |
| --- | --- | --- | --- | --- | --- | --- | --- | --- |
| Sensitivity | 80.56% | 87.67% | 86.54% | 81.38% | 82.90% | 83.84% | 80.07% | 83.28% |
| Specificity | 93.26% | 94.85% | 98.77% | 99.68% | 99.39% | 98.18% | 98.79% | 97.56% |

**Tab 4.** Sensitivity and Specificity results of Contrast effect

**Tab 4.** Sensitivity and Specificity results of Contrast effect
| Method | Backbone | Average Sensitivity | Average Specificity |
| --- | --- | --- | --- |
| GNG-Modified VGG[31] | Resnet-50 | 92.50% | 92.12% |
| Deep Clustering | Resnet-50 | 83.28% | 97.56% |

Fig 3 shows the confusion matrix results for each category, with horizontal and vertical coordinates 1 to 7 denoted as ELBOW, FINGER, FOREARM, HAND, HUMER, SHOULDER, WRIST, respectively. Through the confusion matrix, it is obvious that the results of the FINGER and FOREARM categories are significantly better than the other five categories, achieving good. The results of HUMERUS and SHOULDER are 82.90% and 83.84% respectively. Effective results were also obtained; finally, the results of ELBOW, HAND, WRIST were slightly poor, probably due to the low resolution of the images.

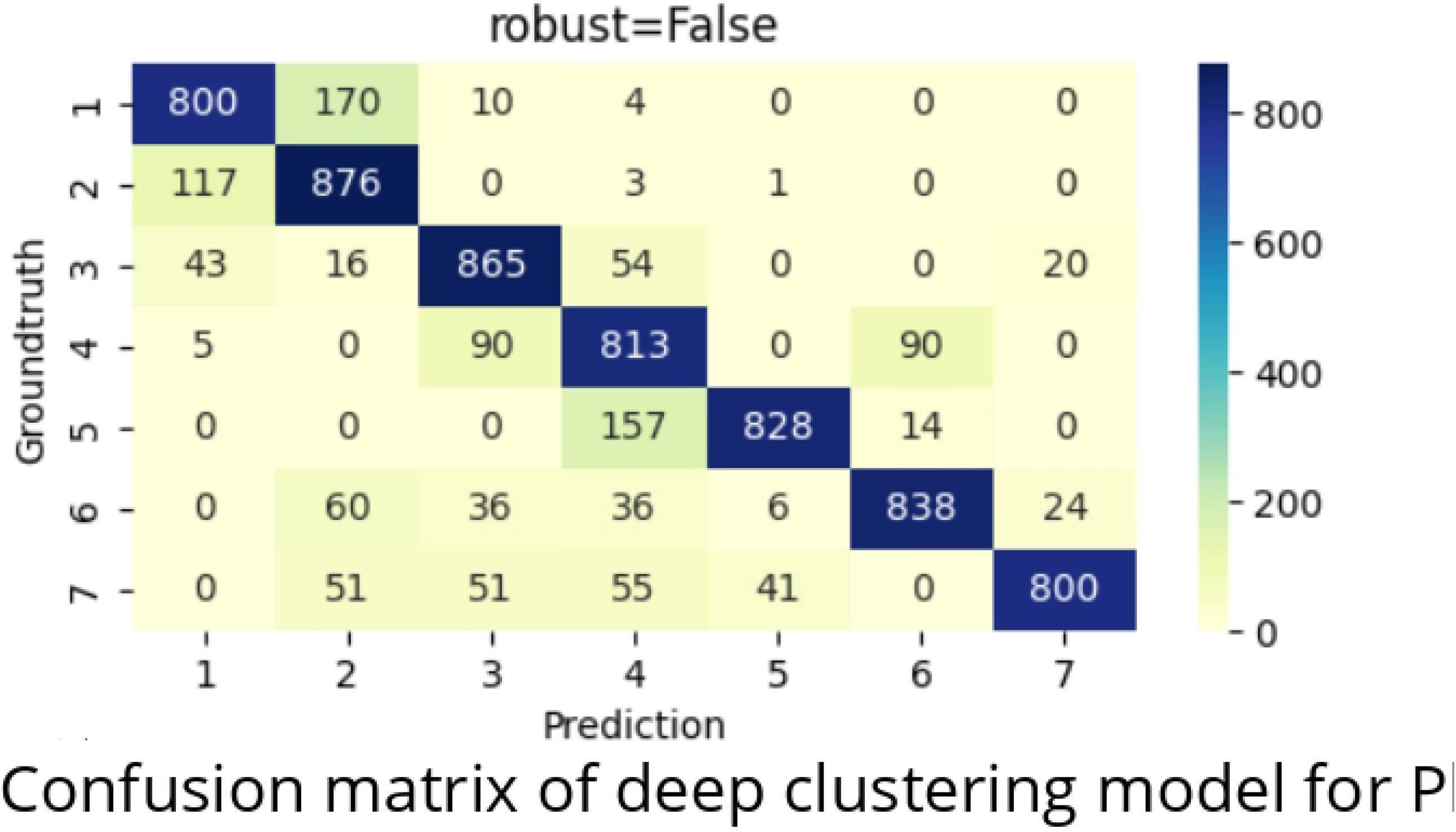

Tab 2 indicates the sensitivity and specificity results of the first stage. The accuracy exceeds 90% for each category of individual usage. Additionally, this phase had the best average sensitivity and specificity at 99.97% and 99.79%, respectively. The second stage’s primary goal was to find any skeletal anomalies. Tab 3 displays the sensitivity and specificity for each bone individually. For this stage, the best mean’s sensitivity and specificity were 83.28% and 97.56%, respectively. It demonstrates the value of the approach suggested in this paper, which can produce accurate classification outcomes while using less training data.

Through comparative analysis, the comparison between the proposed deep clustering and GNG-Modified VGG [31] in this paper yields the results shown in Tab 4. Using the Resnet-50 network architecture and the stage 2 results obtained by unsupervised deep reading clustering, the results are significantly closer than the supervised results. They are able to reduce computation and time consumption while eliminating a large amount of training data, suggesting inspiration for the future use of this method for practical applications.

## 5. Conclusion And Future Work

In this paper, we propose a reliable two-stage unsupervised deep clustering framework detection technique for skeletal anomalies, which combines deep learning to jointly learn the parameters of a neural network together with the clustering assignment of the resultant features. It is trained to detect anomalies. In this paper, experiments were conducted using the MURA dataset, the largest public skeletal image dataset. The first stage’s optimal average sensitivity and specificity were 99.76% and 99.53%, respectively. The optimal average sensitivity and specificity were 83.28% and 97.56%, respectively, for the second stage.

Through extensive experiments on MURA, It is shown that the approach outlined in this work can provide fruitful outcomes and lead to important advances in subsequent research. We anticipate what the future will bring: 1)This paper is limited by the category of the dataset and can only be applied to the x-ray identification of categories and abnormalities of seven upper limb bones. We hope to extend the application to more categories of bone x-ray classification and abnormality detection in the future. 2)There are gaps between datasets, which originate from acquisition devices and regional differences, so this paper hopes to explore some unsupervised frameworks with better generalization performance in the future.

## Data Availability

All relevant data are within the manuscript and its Supporting Information files.

https://stanfordmlgroup.github.io/competitions/mura/

## Acknowledgments

Supported by Jinling Institute of Technology High-level Talent Research Start-up Project(jit-rcyj-202102) ? Key R&D Plan Project of Jiangsu Province (BE2022077) and Jinling Institute of Technology Science and Education Integration Project (2022KJRH18).

## Notes

### Competing Interest Statement

The authors have declared no competing interest.

### Funding Statement

The authors received no specific funding for this work.

### Author Declarations

Ethical approval consent form. This study uses a public database and does not involve ethical issues

## References

1. Fan, L., Li, X., Li, Z., Zhang, Y., Li, J., Li, S., … & Li, B. (2021). A deep learning-based method for bone fracture detection and classification from X-ray images. Journal of medical systems, 45(3), 1–10.

2. Wakefield, Richard J., et al. “The value of sonography in the detection of bone erosions in patients with rheumatoid arthritis: a comparison with conventional radiography.” Arthritis & Rheumatism 43.12 (2000): 2762–2770.

3. Shang, W., Wang, Q., Zhang, L., Jiang, C., & Wu, Z. (2021). A review of machine learning approaches for bone age assessment. Computer methods and programs in biomedicine, 208, 106303.

4. Teven, Chad M., et al. “Fibroblast growth factor (FGF) signaling in development and skeletal diseases.” Genes & diseases 1.2 (2014): 199–213.

5. Celebi, M. Emre, and Kemal Aydin, eds. Unsupervised learning algorithms. Vol. 9. Cham: Springer, 2016.

6. Wu, Xindong, et al. “Data mining with big data.” IEEE transactions on knowledge and data engineering 26.1 (2013): 97–107.

7. De Queiroz, Kevin, and David A. Good. “Phenetic clustering in biology: a critique.” The Quarterly Review of Biology 72.1 (1997): 3–30.

8. McCombie, Gregor, et al. “Spatial and spectral correlations in MALDI mass spectrometry images by clustering and multivariate analysis.” Analytical chemistry 77.19 (2005): 6118–6124.

9. Akçay, H. Gökhan, and Selim Aksoy. “Automatic detection of geospatial objects using multiple hierarchical segmentations.” IEEE transactions on Geoscience and Remote Sensing 46.7 (2008): 2097–2111.

10. Allahyari, Mehdi, et al. “A brief survey of text mining: Classification, clustering and extraction techniques.” arXiv preprint arXiv:1707.02919 (2017).

11. Cheng, David, et al. “A divide-and-merge methodology for clustering.” ACM Transactions on Database Systems (TODS) 31.4 (2006): 1499–1525.

12. Yu, Keping, et al. “Deep-learning-empowered breast cancer auxiliary diagnosis for 5GB remote E-health.” IEEE Wireless Communications 28.3 (2021): 54–61.

13. Zhao, Xiaomei, et al. “A deep learning model integrating FCNNs and CRFs for brain tumor segmentation.” Medical image analysis 43 (2018): 98–111.

14. Torrisi, Mirko, Gianluca Pollastri, and Quan Le. “Deep learning methods in protein structure prediction.” Computational and Structural Biotechnology Journal 18 (2020): 1301–1310.

15. Zhu, Xiaojin, and Andrew B. Goldberg. “Introduction to semi-supervised learning.” Synthesis lectures on artificial intelligence and machine learning 3.1 (2009): 1–130.

16. Dehghani, Mostafa, et al. “Neural ranking models with weak supervision.” Proceedings of the 40th international ACM SIGIR conference on research and development in information retrieval. 2017.

17. Ganin, Yaroslav, and Victor Lempitsky. “Unsupervised domain adaptation by backpropagation.” International conference on machine learning. PMLR, 2015.

18. Li, Shan-Shan. “An improved DBSCAN algorithm based on the neighbor similarity and fast nearest neighbor query.” Ieee Access 8 (2020): 47468–47476.

19. Chen, Yewang, et al. “BLOCK-DBSCAN: Fast clustering for large scale data.” Pattern Recognition 109 (2021): 107624.

20. Likas, Aristidis, Nikos Vlassis, and Jakob J. Verbeek. “The global k-means clustering algorithm.” Pattern recognition 36.2 (2003): 451–461.

21. Lai, Jim ZC, Tsung-Jen Huang, and Yi-Ching Liaw. “A fast k-means clustering algorithm using cluster center displacement.” Pattern Recognition 42.11 (2009): 2551–2556.

22. Peng, Xi, et al. “Deep clustering with sample-assignment invariance prior.” IEEE transactions on neural networks and learning systems 31.11 (2019): 4857–4868.

23. Hershey, John R., et al. “Deep clustering: Discriminative embeddings for segmentation and separation.” 2016 IEEE international conference on acoustics, speech and signal processing (ICASSP). IEEE, 2016.[s2-7] Wen, Jie, et al. “Adaptive graph completion based incomplete multi-view clustering.” IEEE Transactions on Multimedia 23 (2020): 2493–2504.

24. Wen, Jie, et al. “Adaptive graph completion based incomplete multi-view clustering.” IEEE Transactions on Multimedia 23 (2020): 2493–2504.

25. Ghosh, Swarnendu, et al. “Understanding deep learning techniques for image segmentation.” ACM Computing Surveys (CSUR) 52.4 (2019): 1–35.

26. Zhang, Kai, et al. “Beyond a gaussian denoiser: Residual learning of deep cnn for image denoising.” IEEE transactions on image processing 26.7 (2017): 3142–3155.[s2-9] Ghosh, Swarnendu, et al. “Understanding deep learning techniques for image segmentation.” ACM Computing Surveys (CSUR) 52.4 (2019): 1–35.

27. Ren, Yazhou, et al. “Deep density-based image clustering.” Knowledge-Based Systems 197 (2020): 105841.

28. Deng, Zhenyun, et al. “Efficient kNN classification algorithm for big data.” Neurocomputing 195 (2016): 143–148.

29. Li, Xuchun, Lei Wang, and Eric Sung. “Multilabel SVM active learning for image classification.” 2004 International Conference on Image Processing, 2004. ICIP’04. Vol. 4. IEEE, 2004.

30. Sun, Yanan, et al. “Automatically designing CNN architectures using the genetic algorithm for image classification.” IEEE transactions on cybernetics 50.9 (2020): 3840–3854.

31. El-Saadawy, Hadeer, et al. “A Hybrid Two-Stage GNG–Modified VGG Method for Bone X-rays Classification and Abnormality Detection.” IEEE Access 9 (2021): 76649–76661

